# Reliability and Validity of Momentary Pain and Disability Assessments for Lumbar Spine Surgery Patients

**DOI:** 10.1101/2025.09.27.25336802

**Authors:** Saad Javeed, Salim Yakdan, Justin K. Zhang, Kathleen Botterbush, Braeden Benedict, Faraz Arkam, Camilo A. Molina, Brian Neuman, Michael Steinmetz, Michael P. Kelly, Burel R. Goodin, Jay F. Piccirillo, Wilson Z. Ray, Thomas L. Rodebaugh, Madelyn R. Frumkin, Jacob K. Greenberg

**Affiliations:** Department of Neurological Surgery, Washington University, St. Louis, MO, USA; Department of Orthopaedic Surgery, Washington University, St. Louis, MO, USA; Center for Spine Health, Neurologic Institute, Cleveland Clinic, Cleveland, Ohio; Department of Orthopaedic Surgery, Rady Children’s Hospital, San Diego, CA, USA; Department of Anesthesiology, Washington University, St. Louis, MO, USA; Department of Otolaryngology, Washington University, St. Louis, MO, USA; Department of Psychology and Brain Sciences, Washington University, St. Louis, MO, USA; Center for Technology and Behavioral Health; Department of Biomedical Data Science, Dartmouth College, Lebanon, NH, USA

**Keywords:** Back pain, Ecological momentary assessment, Mobile health, Spine surgery, Patient reported outcome measures

## Abstract

**Study Design:** Prospective cohort study.

**Objective:** To evaluate the validity and reliability of ecological momentary assessments (EMAs) of pain and disability in patients undergoing lumbar spine surgery.

**Summary of Background Data:** Patient-reported outcome measures (PROMs) are used to evaluate disease severity and spine surgery outcomes. Traditional cross-sectional PROMs may be limited by recall-bias and inability to capture longitudinal symptom dynamics. Momentary symptom evaluations using brief mobile surveys (i.e., EMAs) can overcome these limitations by capturing symptom severity and its change over time in patients’ natural environments.

**Methods:** Adults undergoing lumbar/thoracolumbar spine surgery completed EMAs five-times daily for approximately 3-weeks preoperatively to assess momentary pain and disability. Participants also completed traditional PROMs, including Patient-Reported Outcomes Measurement Information System (PROMIS) pain-intensity, pain-interference, and Oswestry Disability Index (ODI). Several analyses were performed to evaluate reliability of EMA items, composites, and summary metrics, their variability within-patients over time, construct-validity, and convergent-validity.

**Results:** A total of 123 patients, 46% males, and mean age 57-years (+/-13) were enrolled. EMA metrics demonstrated high item-reliability, composite-reliability, and temporal-stability of assessments over time. In validity analysis, EMA pain and disability showed strong correlation with PROMs, including PROMIS pain intensity, pain interference, and ODI. Nonetheless, there was substantial variability of EMAs in each severity category of PROMs. For example, among patients with high PROMIS pain-intensity, the average EMA pain score was 70 but ranged from 22 to 97 (+/-20). Several variables including sex, age, and ODI were associated with EMA symptom variability. In multivariable analysis, EMA pain and disability (mean, variability, and mean*variability interaction) explained substantial variability in PROMs.

**Conclusion:** Momentary assessments of symptom severity using EMAs is a valid and reliable approach to evaluate spine-related pain and disability in everyday life.

## Introduction

Degenerative spine disease is one of the most common public health problems that results in debilitating chronic back and leg pain.^1,2^ Although spine surgery is only reserved for a subset of patients not responding to conservative treatments, in the US alone, nearly 400,000 spine fusion surgeries are performed annually to treat degenerative spine disease.^3–5^

Pain and spine-related disability are the most common complaints in patients suffering with degenerative spine disease.^6^ To evaluate these symptoms, various patient reported outcome measures (PROMs) are used to quantify disease severity and evaluate postoperative outcomes.^7– 13^ Traditional PROMs are administered infrequently and ask patients to summarize their experiences over the recent past (e.g., past week).^14–16^ This design increases the likelihood of recall bias and limits the ability of traditional PROMs to assess longitudinal symptom dynamics that may provide unique insights into spine disease.^16–18^

Mobile health (mHealth) technology offers the ability to overcome these shortcomings and evaluate symptom severity in everyday life. In particular, ecological momentary assessment (EMA) uses brief mobile surveys, typically delivered via smartphone, to evaluate momentary symptom severity, often multiple times daily.^19–21^ While such methods have been reported in spine surgery patients, given their novelty, EMA measures of pain and disability have not been validated in lumbar disease populations.^22,23^ Therefore, the objective of this study was to evaluate the reliability and validity of pain and disability EMA in patients with degenerative lumbar spine disease.

## Materials and Methods

### Study Design and Participants

We enrolled patients undergoing elective lumbar and thoracolumbar surgery for degenerative spine disease at a single institution between February 2021 to March 2023. Inclusion criteria included adult patients 21-85 years of age who owned a smartphone, had at least one week to complete preoperative assessments, and had a numeric rating scale score of at least 3/10 back and/or leg pain in the week prior to enrollment.^23^ Patients undergoing isolated thoracic surgery, surgery for malignancy, trauma, and infection, or those undergoing another major surgery within 3 months of data collection were excluded. Written informed consent was obtained from all study participants. Ethics approval was obtained from the institutional review board at our institution (#202012139).

### mHealth Protocol

After enrollment, participants downloaded the LifeData application (LifeData LLC.) and selected a preferred time window to receive 5 daily EMAs, delivered 3 hours apart. The preoperative mHealth evaluations lasted approximately 3 weeks, depending on the time until surgery. The mHealh protocol was described in detail previously.^23^ In addition to mHealth evaluations, participants completed traditional PROMs, including the Oswestry Disability Index (ODI) and Patient Reported Outcomes Measurement Information System (PROMIS) pain-interference, pain-intensity, and numerical rating scale (NRS) back pain and leg pain prior to surgery.

### EMA Measures

At EMA administration, three items related to pain severity (overall pain, leg pain, and back pain) and pain-related disability (interference with activity, concentration, and enjoyment) were assessed on a scale from 0 (“Not at all”) to 100 (“Worst possible”). These items were selected based on expert opinion and their similarity to established PROMs.^11^ (**etable 1**) The composites were then pooled into daily person-level summary indices of *Pain* and *Interference*, as shown in **Figure 1**.

**Figure 1:**
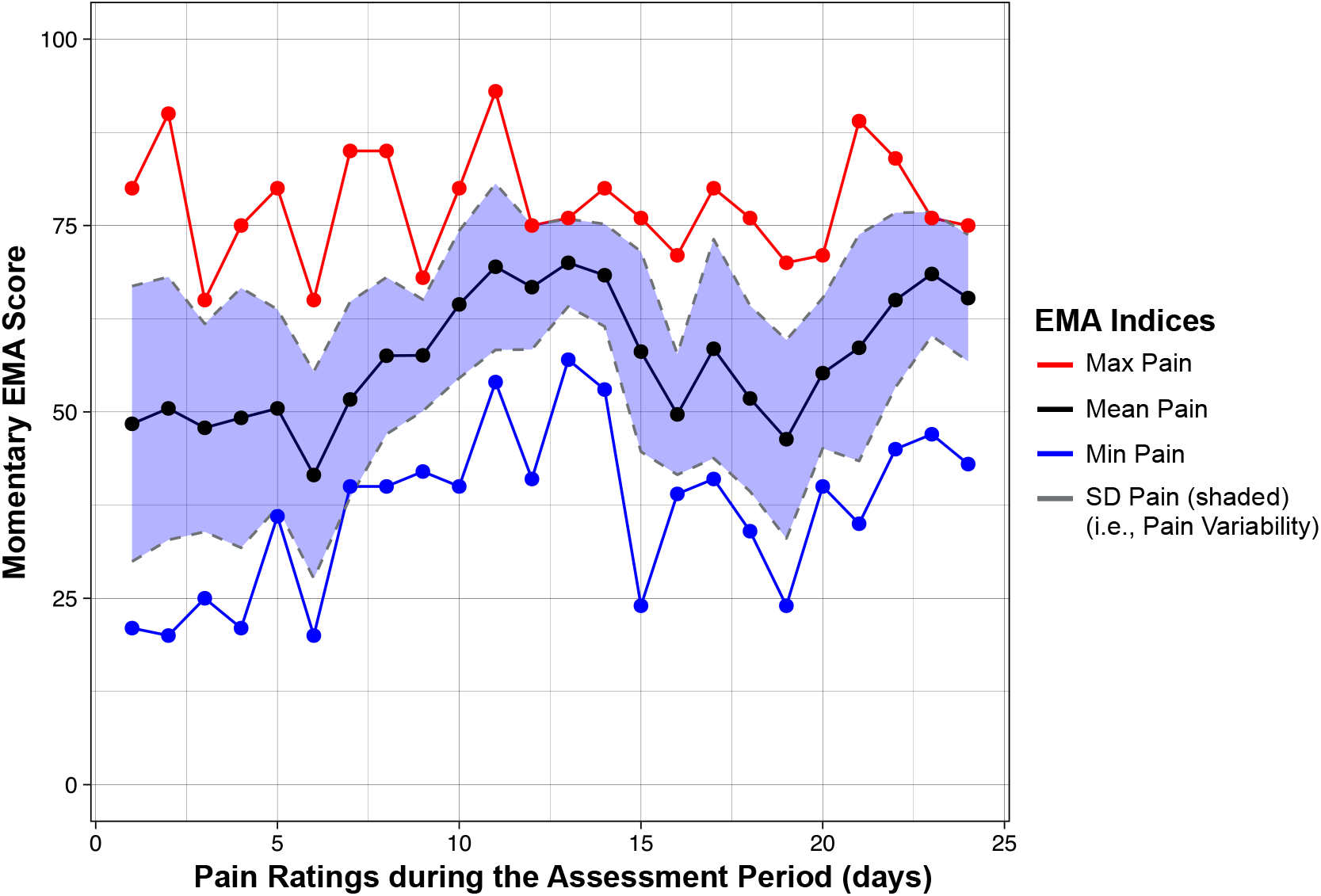
Summary Indices of EMA in an example participant completed during the assessment period. The max pain was defined as maximum pain rated on a day, the min pain was defined as minimum pain rated on a day, mean pain was calculated by taking average of pain ratings throughout the day. And finally, pain variability indicated the standard deviation of pain ratings throughout the day.

### Statistical Analyses

#### Item Reliability

Given that EMA are collected repeatedly from individuals over time, observations are not independent. Pooling all data to calculate reliability metrics (e.g., Cronbach’s alpha) therefore violates important assumptions and can lead to inaccurate estimates of reliability.^24,25^ We therefore used procedures described by Geldhof and colleagues^26^ to estimate multilevel omega (ω) coefficients based on multilevel confirmatory factor analysis. Within-person reliability (ω_within_) reflects internal consistency of the three items comprising each composite score (analogous to Cronbach’s alpha). Between-person reliability (ω_between_) reflects the degree to which responses differentiate between individuals.

#### Composite Reliability

We also calculated several reliability metrics based on *pain* and *interference* composite scores (created by averaging the three relevant items for each individual at each time point). The intraclass correlation coefficient (ICC) was calculated to quantify the ratio of variance between vs. within individuals. Similarly, *between-person reliability* was defined as the ratio of composite EMA score variance between participants to overall variance, controlling for the average number of observations per participant.^27,28^ In addition, since the duration and fidelity of EMA completion varies, the impact of *EMA density* on ICC and between-person reliability was assessed by randomly sampling varying number of observations and re-calculating both metrics.^25,28^

#### Summary Metric Reliability

Finally, we examined split-half reliability of person-level summary metrics commonly derived from EMA data. For example, *average severity* was calculated by taking the average of all EMA pain severity ratings during the assessment period for each individual. Symptom *variability* was based on the within-person standard deviation in each domain, a measure which has been suggested to be associated with greater emotional distress and functional limitations.^29,30^ To estimate the reliability of these person-level metrics, we calculated split-half reliability by splitting each individual’s data into two halves (odd and even surveys), calculating the mean and SD for each individual in each of the split data sets, and quantifying the correlation between the split-halves using Spearmen-Brown correlation coefficients.^31,32^

#### Concurrent Validity

To evaluate the validity of EMA summary metrics, we assessed convergent validity by examining correlations between each *pain* and *interference* summary index and three PROMs: ODI, PROMIS pain-interference, and pain-intensity. Our hypothesis was that there would be a moderate to strong correlation between EMA and PROMs, indicating EMA and PROMs captured similar but not identical information. We also graphed mean EMA pain and disability scores, stratified by traditional PROM severity values, to visualize variability within each category. To further evaluate convergent validity, we examined the correlation of individual EMA pain items (overall pain, back pain, and leg pain) with retrospective NRS back and leg pain scores.

#### Predictors of High Symptom Variability

In patients with highly variable symptom severity, EMA assessments can reveal unique patterns not captured by traditional PROMs. However, EMAs may have less value in patients with less symptom variability. For this reason, we aimed to identify a subset of patients in whom EMA assessment are more variable. To identify patients with highly variable symptoms, we used linear regression analysis with symptom variability (i.e., pain and interference standard deviation) as the outcome. An exhaustive model search identified variables linked to symptom variability, and the final variables were selected based on the model with lowest Akaike Information Criteria (AIC).

#### Predictors of PROMs

Finally, we were interested in examining the degree to which recalled measures of pain/disability reflected average symptoms versus symptom variability captured with EMA. PROMs based on retrospective recall (e.g., ODI, PROMIS, NRS) are intended to capture average or typical levels of pain and disability. However, prior EMA research has demonstrated that individuals’ most severe and most recent pain experiences influence recall of average pain related to rheumatoid arthritis and knee osteoarthritis.^33–35^ In a small study of patients with low back pain, there was less evidence of recall bias when comparing recalled weekly pain intensity to daily assessments.^36^ In the current study, we used linear regression to assess the relative contributions of average symptom severity and symptom variability to retrospective PROMs. We further explored the possibility of an interaction between average symptom severity and symptom variability in predicting PROMs. A significant interaction would suggest that symptom variability may have a different impact on recalled symptom severity at different average pain/disability levels.

#### Statistical Analysis

Frequencies and proportions for categorical variables and mean (SD) or median (IQR) for continuous variables were used as appropriate. The Pearson correlation coefficients (r) were calculated for correlation analyses between EMAs and PROMs. The correlation coefficients were considered weak if r < 0.3, moderate if r ≥ 0.3 and r < 0.5, and strong if r ≥ 0.5.^37^ Since several EMA metrics had non-normal distribution, non-parametric statistics were used to compare differences. Multivariable linear regression analysis was used to analyze the criterion validity of EMA measures and association with PROMs. For linear regression analysis, all EMA measures were normalized by scaling and centering the variables. The R^2^ values of linear models for each subset of EMA measures were quantified to evaluate the magnitude of association. The continuous-to-continuous interactions were evaluated by graphing change of interacting variable (*variability*) with a moderator variable (*mean*) held constant at several incremental values.^38^ The threshold for significance was set at 2-tailed α < 0.05. All analyses were performed in R version 4.2.1 (R Project for Statistical Computing). Exhaustive model search was performed using glmuti version 1.0.8.

## Results

### Cohort Characteristics

Between 2021 to 2023, a total of 123 patients were enrolled and completed the preoperative mHealth protocol. Patients had a mean age of 58 (+/-13) years, with 56 (46%) being male. The median number of spinal fusion levels was 2 (range: 1-19). A detailed description of participant characteristics is given in **Table 1**.

**Table 1:** Participant Characteristics.

| **Table 1: Participant Characteristics** |  |
| --- | --- |
| **Characteristic** | **% (N) of participants (N=123)** |
| Age, mean (SD), y | 59 (13) |
| **Sex** |  |
| Male | 46 (57) |
| Female | 54 (66) |
| BMI, mean (SD), y | 29 (5) |
| **Race** |  |
| White | 95 (117) |
| Black | 3 (4) |
| Other | 2 (2) |
| **Ethnicity** |  |
| Non-Hispanic | 96 (117) |
| Hispanic | 4 (5) |
| **Employment Status** |  |
| Actively working | 36 (44) |
| Homemaker | 5 (6) |
| Unemployed | 2 (3) |
| Retired | 37 (45) |
| On disability | 20 (25) |
| **Education Level** |  |
| Did not complete high school | 3 (4) |
| High school degree | 35 (43) |
| College degree | 33 (41) |
| Graduate degree | 29 (35) |
| **Spine specific pattern of symptoms** |  |
| Back pain | 57 (70) |
| Leg pain | 56 (69) |
| Neurogenic claudication | 19 (24) |
| Motor deficit | 9 (12) |
| Sensory deficit | 15 (19) |
| **Surgical Approach** |  |
| Levels of surgery median (range) | 2 (1-19) |
| Posterior | 62 (76) |
| Anterior | 9 (11) |
| Anterior/posterior | 27 (33) |
| Not performed/postponed at time of study | 2 (3) |

### Reliability of EMAs

Detailed reliability results are provided in **Table 2**. The *pain* and *interference* items showed good within- and between-person reliability (ωs > 0.78). The ICCs for the pain and interference composite scores were 0.77 and 0.83, respectively, suggesting that 77-83% of the variance occurs between persons. Adjusting for the average number of assessments (*T* = 76), both composite scores also showed excellent between-person reliability. ICC and between-person reliability were not sensitive to EMA density (i.e., number of randomly sampled observations), and between-person reliability was excellent (>0.9) with as few as 5 EMAs **(Figure 2)**. Both composites also had excellent split-half reliability for both means and standard deviations (all coefficients ≥0.96, **Table 2**).

**Table 2:** Reliability of EMA Measures.

| Table 2: Reliability of EMA Measures |  |  |  |
| --- | --- | --- | --- |
| Item reliability<sup>a</sup> |  |  |  |
| Items | Omega<sup>b</sup> within-person | Omega<sup>b</sup> between-person |  |
| Pain Items |  |  |  |
| Overall pain,<br>back pain,<br>leg pain | 0.84<br>95% CI (0.83-0.85) | 0.85<br>95% CI (0.81-0.90) |  |
| Interference Items |  |  |  |
| Enjoyment,<br>activities,<br>concentrate | 0.78<br>95% CI (0.77-0.79) | 0.94<br>95% CI (0.92-0.96) |  |
| Composite reliability<sup>c</sup> |  |  |  |
| Composites | Within-person (ICC) | Between-person (ICC) |  |
| Pain | 0.77 | 0.99 |  |
| Interference | 0.83 | 0.99 |  |
| Composite reliability (split sample in even and odd surveys) |  |  |  |
| Composites | Pearson brown Correlation coefficients<sup>d</sup> |  |  |
|  | Mean | Standard deviation | Standard deviation/<br>mean ratio |
| Pain | 0.99 | 0.97 | 0.98 |
| Interference | 0.99 | 0.96 | 0.99 |
| <sup>a</sup> Reliability based on each item in each survey per participant during the assessment period. |  |  |  |
| <sup>b</sup> Omega coefficients quantify the reliability of overall scale and reliability within each subgroup or levels. |  |  |  |
| <sup>c</sup> Reliability of composite mean of each item in each survey per participant during the assessment period. |  |  |  |
| <sup>d</sup> Spearman correlation between even and odd surveys per participant, |  |  |  |
| Within-person: consistency of measures in same participant over multiple time-points. |  |  |  |
| Between-person: consistency of measures over different participants. |  |  |  |

**Figure 2:**
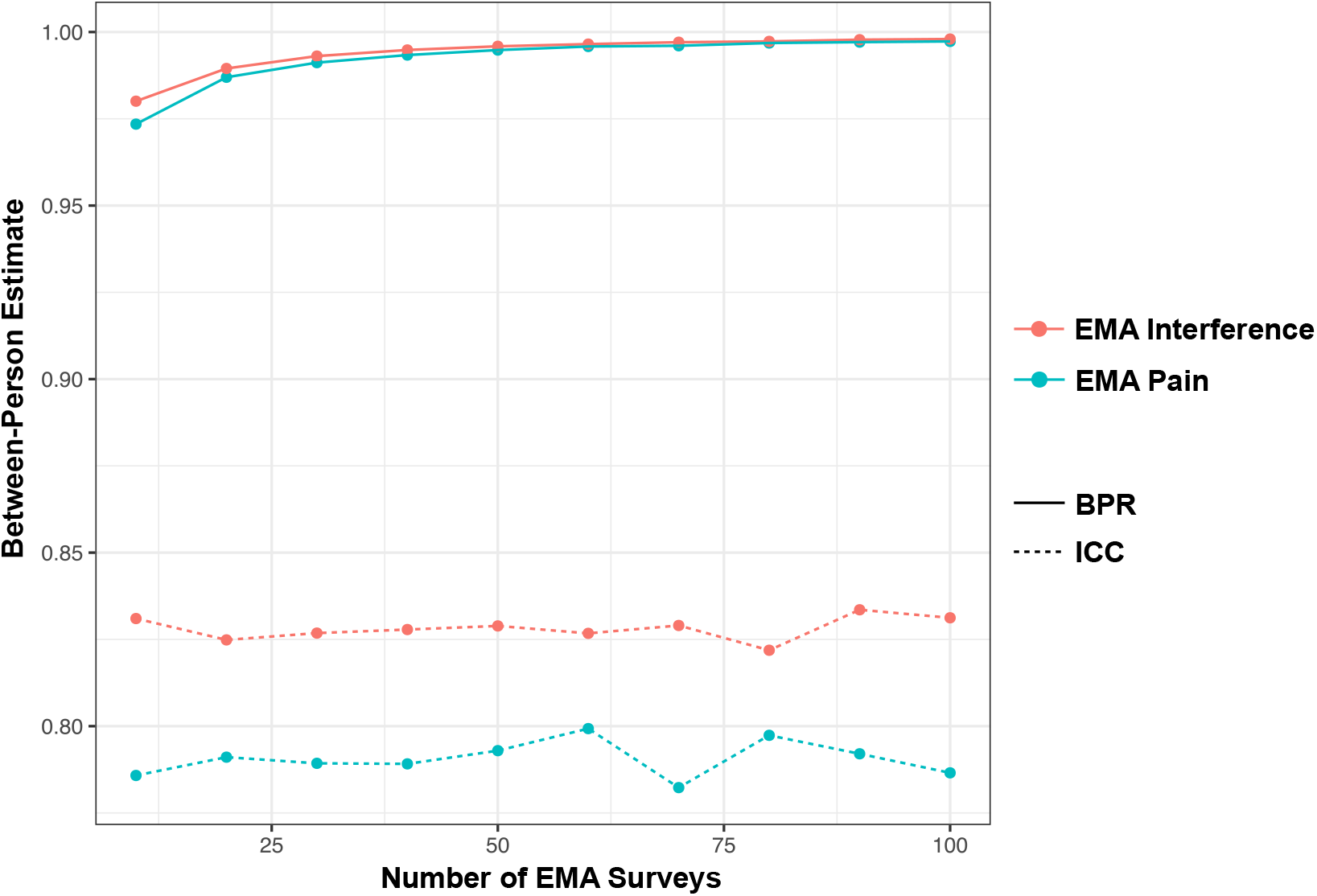
Reliability of EMA pain and interference by varying density of assessments. The solid line indicates between-person reliability (BPR), and dotted line indicates intra-class correlation coefficient (ICC).

### Convergent Validity of EMA Measures and Correlation with PROMs

Among the average EMA measures, there was a significant correlation between mean-pain and PROMIS pain-intensity, (r= 0.66, 95% CI: 0.54-0.75, p<0.001), mean-interference and PROMIS pain-interference, (r= 0.64, 95% CI: 0.53-0.74, p<0.001), and mean-interference and ODI (r= 0.65, 95% CI: 0.54-0.74, p<0.001). The correlation analyses of EMA measures over the assessment period with PROMs is shown in **Figure 3**.

**Figure 3:**
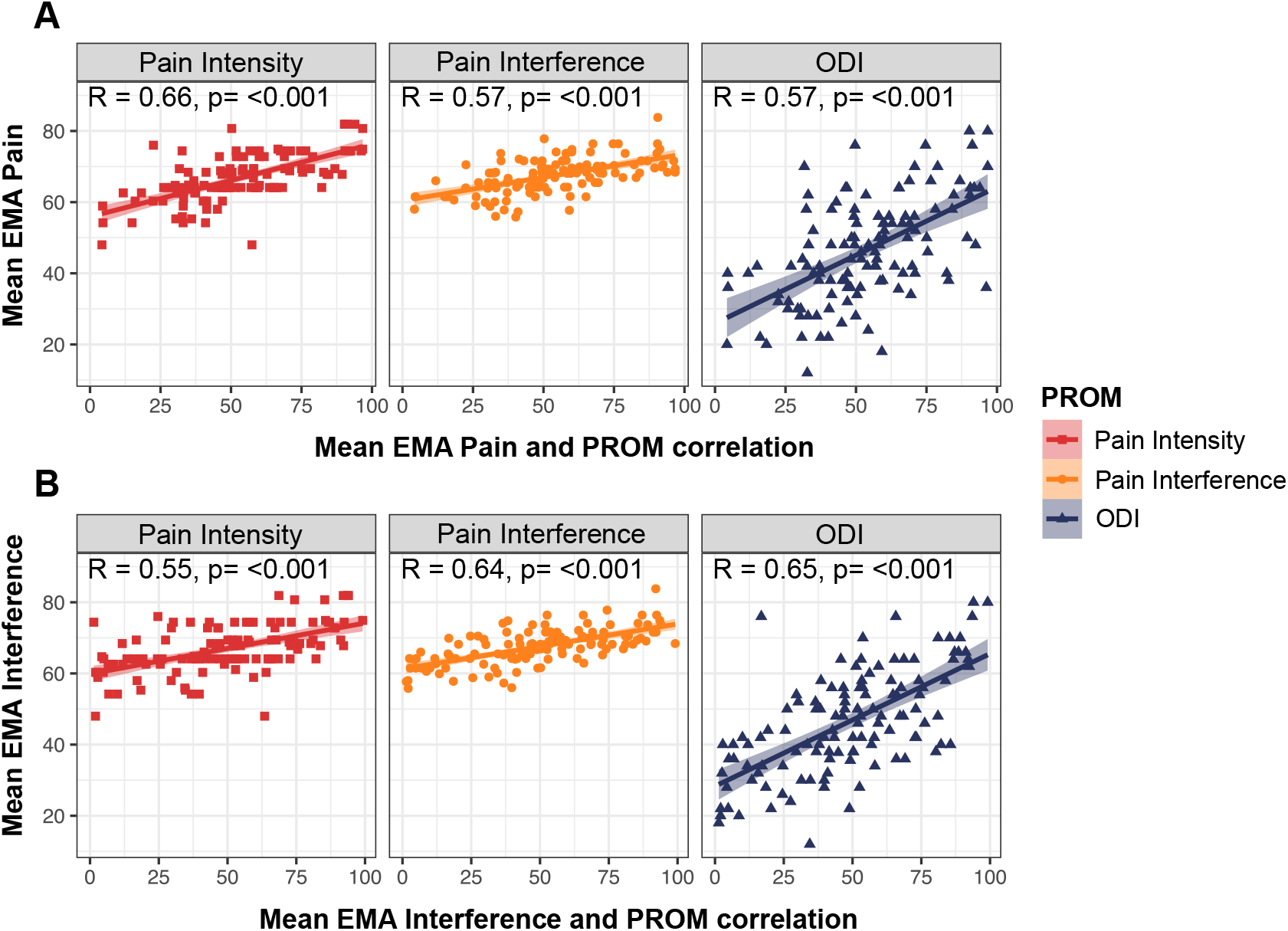
Correlation of mean EMA pain and interference during the assessment period with traditional patient reported outcome measures (PROMs). **A)** Correlation of mean EMA pain with PROMIS pain intensity, pain interference, and Oswestry disability index (ODI). **B)** Correlation of mean EMA interferences with PROMIS pain intensity, pain interference, and ODI). R indicates Pearson correlation coefficients with p value. Each point indicates specific PROM, line indicates linear correlation coefficient, and shaded area around the correlation indicates 95% confidence interval of correlation.

The mean-pain and mean-interference sub-grouped by severity groups of ODI, PROMIS pain-interference and pain-intensity is shown in **Figure 4**. There were significant differences in the mean-pain and mean-interference in low, medium, and high severity categories of PROMIS pain-intensity, pain-interference, and ODI (all p<0.001). Although low PROMIS pain-intensity was associated with lower average EMA-pain than medium and high PROMIS pain-intensity (29 vs. 51 and 70), there was substantial variability within each group. For example, the high PROMIS pain-intensity group had standard-deviation of 20, with mean EMA pain scores ranging from 22 to 97 **Figure 4**

**Figure 4:**
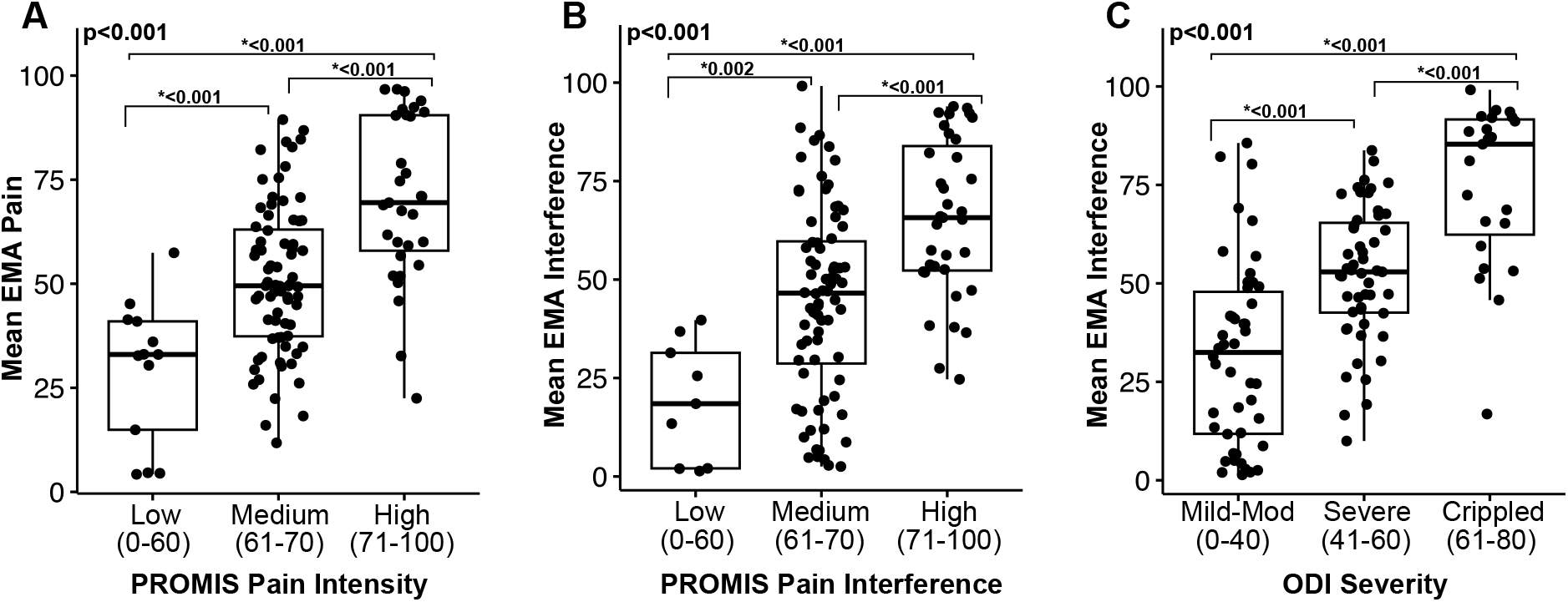
The mean EMA pain and interference in severity categories of **A)** PROMIS pain intensity, **B)** PROMIS pain interference, and **C)** Oswestry disability index (ODI). ODI severity categorized as minimal disability (ODI=0-20), moderate disability (ODI=21-40), and severe disability (ODI=41-60), and crippled/bed-bound (ODI=61-100).^41^ The PROMIS pain interference and pain intensity categorized as mild (t-score=55-60), moderate (t-score=60-70), and severe (t-score=70-100).^42^ The bold p-value indicates Kruskal-Wallis one way analysis of variance statistic and p-values in asterisk indicate pairwise comparisons of each group.

As shown in **Figure 5A**, retrospective NRS back pain was strongly positively correlated with average EMA back pain (r=0.68, p<0.001) and average EMA overall pain (r=0.64, p<0.001). By contrast, retrospective NRS leg pain was strongly positively correlated with average EMA leg pain (r=0.76, p<0.001), but only weakly associated with average EMA overall pain (r=0.28, p<0.001, **Figure 5B**).

**Figure 5:**
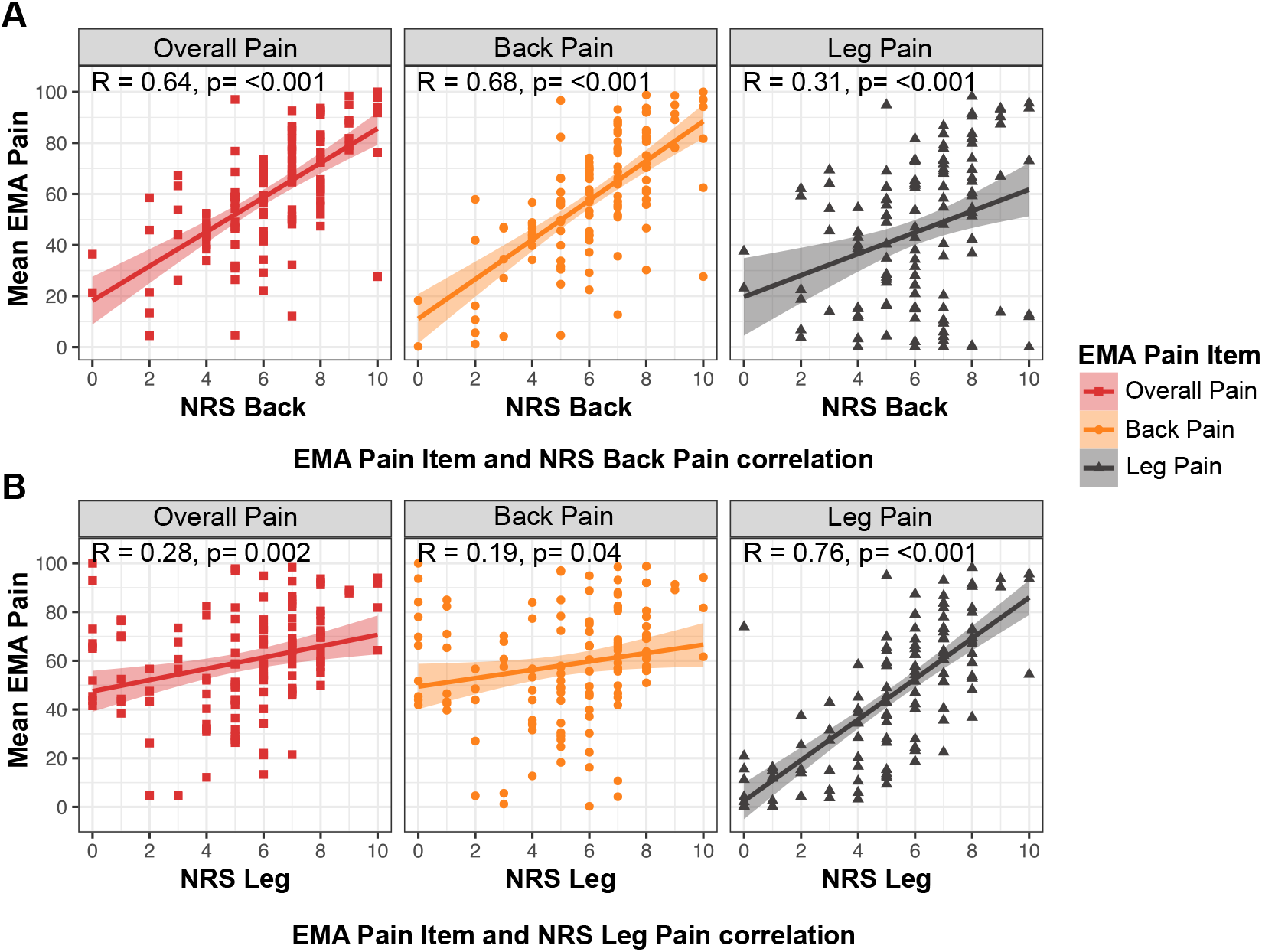
Correlation of mean EMA pain items and traditional patient reported numeric rating scale (NRS) back and leg pain. **A)** Correlation of mean EMA pain items with NRS *back pain* scores. **B)** Correlation of mean EMA pain items with NRS *leg pain* scores. R indicates Pearson correlation coefficients with p value. Each point indicates specific EMA pain item, line indicates linear correlation coefficient, and shaded area around the correlation indicates 95% confidence interval of correlation.

### Prediction of Symptom Variability

Several candidate variables were tested for their association with pain and interference variability. Among all the variables, sex, ODI, and levels-fused were top three predictors of EMA pain variability **(eFigure 1A; eTable 2)**. Similarly, sex, age, PROMIS pain-interference, and ODI were top four predictors of EMA interference variability **(eFigure 1B; eTable 2)**.

### EMAs incremental association with PROMs

The multivariable linear regression analyses examined whether different EMA summary metrics explained unique variance in recalled PROMs. The association of mean and variability of EMA measures with PROMs is shown in **Figure 6 and eFigure 2**. A combination of mean and variability of EMA back and leg pain specific items, and interaction between these two variables explained substantial (51-61%) variability in the NRS back and leg pain (p<0.001). Specifically, there was a significant interaction such that at *higher* pain severity levels, greater variability in leg/back pain was associated with *lower* NRS back and leg pain; (interaction of mean and variability back pain on NRS back pain: β=-0.35, p=0.01; and between mean and variability leg pain on NRS leg pain: β=-0.38, p=0.02 **(Figure 6)**.

**Figure 6:**
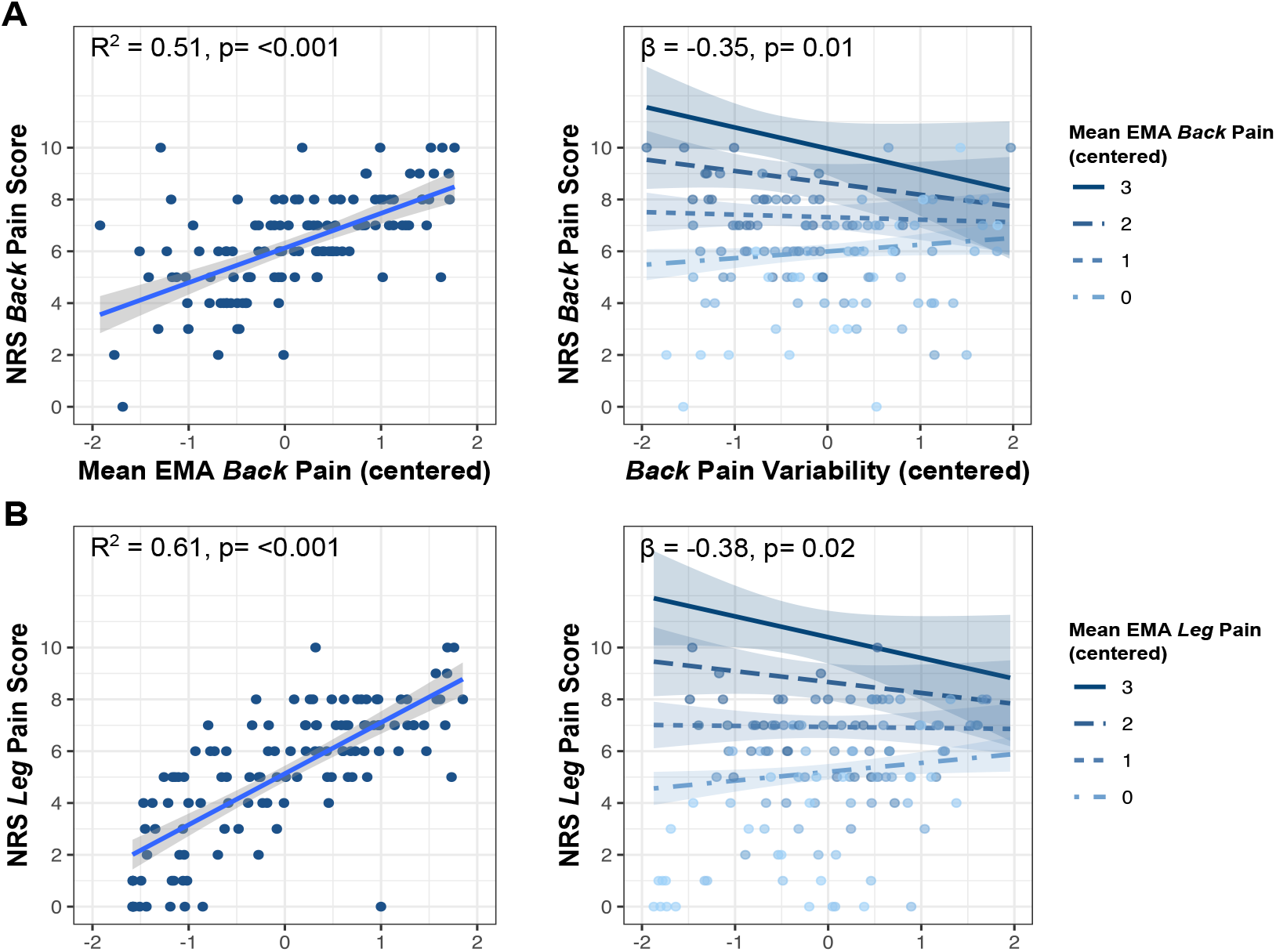
Incremental association of EMA pain items summary indices with traditional patient reported numeric rating scale (NRS) back and leg pain. **A)** Association of mean EMA *back pain* with NRS *back pain* and interaction of *back pain* variability with mean EMA *back pain* moderating at various levels. **B)** Association of mean EMA *leg pain* with NRS *leg pain* and interaction of *leg pain* variability with mean EMA *leg pain* moderating at various levels. The R^2^ indicates the variability explained by the mean, variability, and interaction of mean and variability of each of EMA pain item. The β indicates the regression estimate of the interaction. **Legend:** A value of 0 represents the mean of the centered variable, while 1, 2, and 3 correspond to points above the average.

Similarly, at higher levels of mean EMA-pain and EMA-disability, more symptom variability was associated with lower PROMIS Pain Intensity (β=-0.76, p=0.11) and ODI severity (β=-1.85, p=0.15), respectively. **(eFigure 2)**. However, these interaction effects were not significant.

## Discussion

In this prospective study, we established the reliability and validity of EMA-based pain and disability measures for lumbar spine surgery. Our analysis found that momentary assessment using EMA is a reliable method to evaluate spine-related pain and disability. EMA-based momentary measures of pain and disability show convergent validity with PROMs but offer unique insights compared to traditional PROMs. Our analysis also found that distinct indices of momentary assessments, including mean and variability, capture independent aspects of recalled pain and interference, providing new insights into spine disease-related disability. These results lay the foundation for broader adoption of EMA in spine surgery, enabling researchers and clinicians to capture variability in symptoms over time.

In the era of personalized medicine, there has been considerable interest in more individualized approaches to treating degenerative spine disease.^39–41^ Currently, disease severity is evaluated using validated PROMs at intermittent clinical visits. Although meaningful, these PROMs have important limitations. They are typically administered once or a few times in the preoperative and postoperative settings, increasing risk of recall bias.^36,42^ Our findings add to previous concerns of recall bias using traditional PROMs to assess pain, as symptom variability explained unique variance in PROMs when accounting for the average of EMAs. Recent prediction models for surgical outcomes in large datasets have shown relatively poor reproducibility in external validation, potentially reflecting, in part, the lack of precision in the PROMs used to define disease severity.^43^ Improved measurement of disease severity may facilitate improved predictions of disease outcomes for patients with spine disease.

Through real-time capture of patient-reported symptoms, EMAs also provide opportunities to develop novel features to improve prediction of treatment outcomes. For example, EMAs can be used to examine dynamic relationships between pain and psychosocial symptoms. EMAs can be further combined with passive physiological monitoring to examine person-specific relationships between patient-reported symptoms and activity, sleep, or physiological arousal. ^22,23,30,39,44^ Observing within-person associations patient-reported symptoms requires symptom variability (e.g., pain fluctuations). Our analyses demonstrated that sex, age, and spine disease related symptoms severity predicted symptom variability. These results may inform future research in designing customized EMA approaches to evaluate disease severity in spine surgery.

Though potentially showing a clinically meaningful interaction with symptom severity, symptom variability on its own had a fairly weak association with recalled measures of pain and disability. This finding is consistent with recent studies exploring symptom variability in the chronic low back pain literature.^16,29,30^ This result suggests that symptom variability on its own, without considering symptom severity, may not have a strong impact on recalled symptom severity. However, to what extent symptom variability impacts surgical outcome remains unknown.^16,30^ Therefore, future studies will be needed to examine how symptom variability profiles might influence response to surgical treatment.

Beyond their association with various PROMs, effective use of EMAs depends on the reliability of the assessments. Our reliability analyses revealed that both pain and disability EMAs exhibited high item and composite reliability. Furthermore, reliability remained stable across varying numbers of observations and temporal position within the assessment window. These findings demonstrate the feasibility of using EMAs in clinical settings, where strict adherence to survey protocols may not be practical for many patients.

### Limitations

This study has several important strengths including a prospective study design, and rigorous analytical methods for validation and reliability analyses. Nevertheless, this study has several limitations. First, this was a single center study including a relatively small patient cohort limiting generalizability of our findings. Second, our study population was predominantly White and with at least high school degree, emphasizing the importance of validation in more diverse population. Finally, although this study included several summary indices derived from EMAs, we did not explore a variety of other complex longitudinal measures (*e*.*g*., autocorrelation, multimodal associations). Though requiring more advanced statistical techniques to derive, such EMA-based measures may offer additional insights that should be explored in future studies^16,45,46^.

## Conclusions

In this prospective study, we established the reliability and validity of momentary pain and disability assessments in patients undergoing lumbar spine surgery. Our findings demonstrate the suitability of using mobile health technology for momentary assessments related to degenerative lumbar spine disease. This approach holds substantial future promise for capturing personalized disease profiles and supporting individualized treatment strategies for spine disease.

## Data Availability

All data produced in the present study are available upon reasonable request to the authors

