## Supplemental File for "Reliability and Validity of Momentary Pain and Disability Assessments for Lumbar Spine Surgery Patients"

### eTable 1: Preoperative EMA items included for patients undergoing lumbar/thoracolumbar surgery.

**EMA Items**

Please rate each item on a scale from Not at all (0) to Worst possible (100).

1. Right now, how intense is your back pain?
2. Right now, how intense is your leg pain?
3. Right now, how intense is your overall pain?
4. Right now, how much is pain interfering with your enjoyment of life?
5. Right now, how much is pain interfering with your activities?
6. Right now, how much is pain interfering with your ability to concentrate?

| eTable 2: Multivariable regression of variables predicting EMA symptom variability | | | | | |
| --- | --- | --- | --- | --- | --- |
| **Pain Variability (SD)** | | | **Interference Variability (SD)** | | |
| **Variables^a^** | **Beta (95% CI)** | **p value** | **Variables^a^** | **Beta (95% CI)** | **p value** |
| **Sex female** | **0.56 (0.16 to 0.91)** | **0.005** | Sex female | 0.34 (-0.02 to 0.70) | 0.06 |
| No. of levels fused | -0.03 (-0.07 to 0.01) | 0.17 | **Age** | **-0.02 (-0.03 to -0.003)** | **0.01** |
| ODI | -0.008 (-0.02 to 0.005) | 0.21 | **PROMIS pain-interference** | **0.06 (0.02 to 0.11)** | **0.008** |
|  |  |  | **ODI** | **-0.02 (-0.03 to -0.001)** | **0.03** |
| ^a^Variables were selected from model with lowest Akaike Information Criterion (AIC) | | | | | |

### **eFigure 1:** Variables Predicting EMA pain variability and interference variability.

(B)

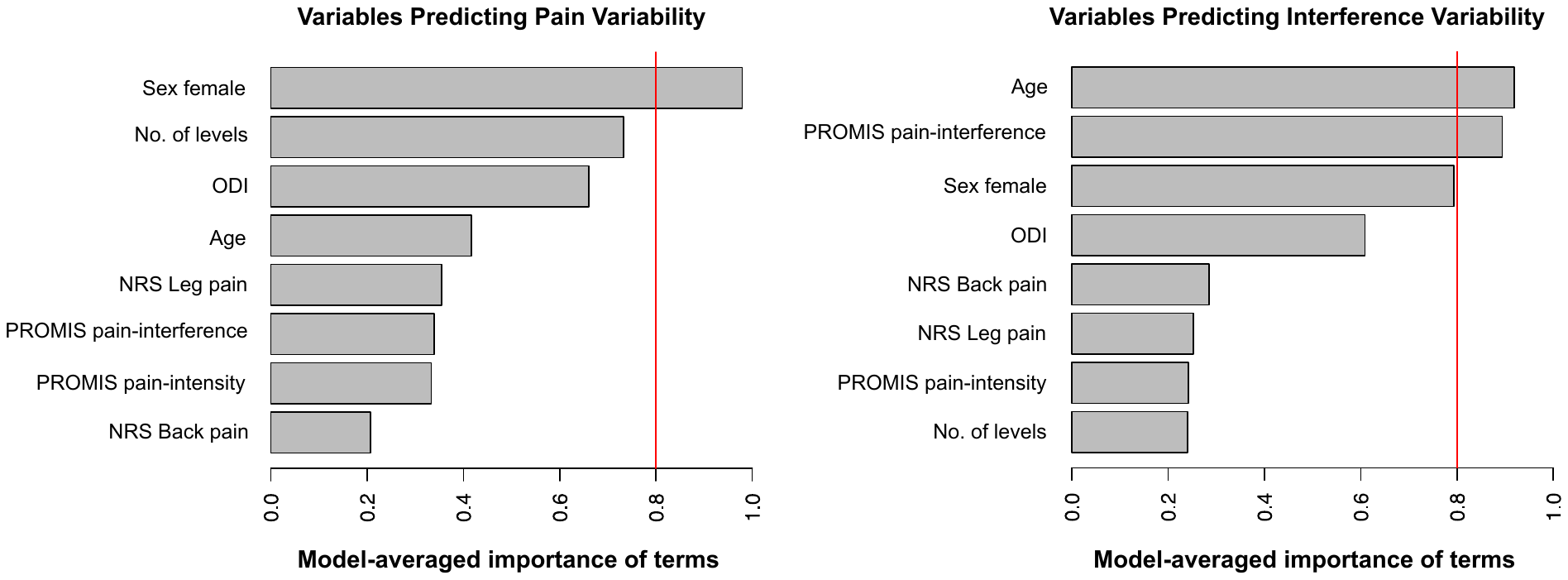

(A)

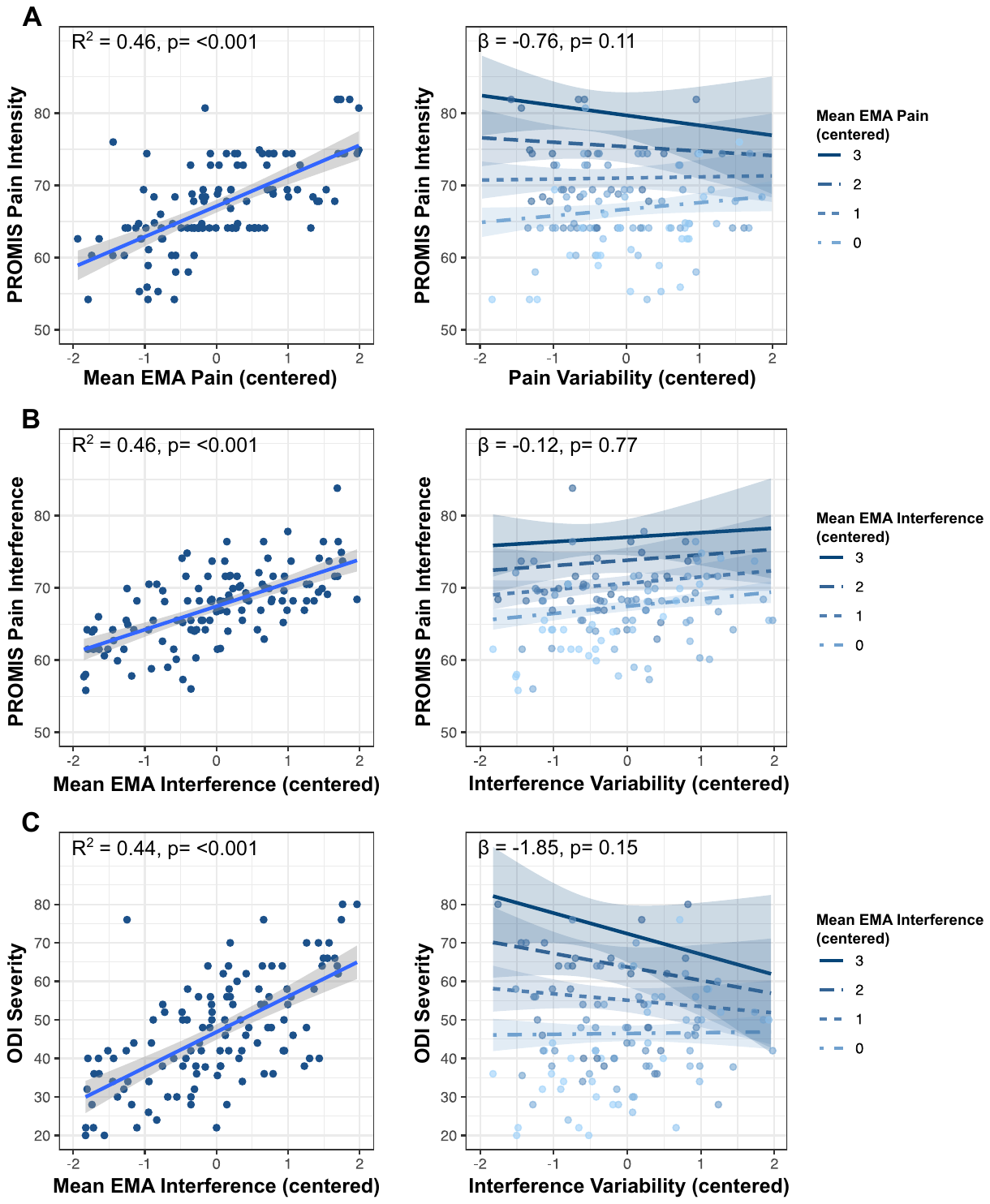
**eFigure 2:** Incremental association of EMA summary indices with traditional patient reported outcome measures (PROMs). **A)** Association of mean EMA pain with PROMIS pain intensity and interaction of pain variability with mean EMA pain moderating at various levels. **B)** Association of mean EMA interference with PROMIS pain interference and interaction of interference variability with mean EMA interference moderating at various levels. **C)** Association of mean EMA interference with Oswestry disability index (ODI) and interaction of interference variability with mean EMA interference moderating at various levels. The R^2^ indicates the variability explained by the mean, variability, and interaction of mean and variability of each of EMA pain and interference. The β indicates the regression estimate of the interaction.
